# Knowledge, Attitudes, and Practices (KAP) Towards COVID-19: An Online Cross-Sectional Survey of Tanzanian Residents

**DOI:** 10.1101/2020.04.26.20080820

**Authors:** Sima Rugarabamu, Mariam Ibrahim, Aisha Byanaku

**Affiliations:** Department of Oral health, Muhimbili National Hospital-Mloganzila, Dar-Salaam, Tanzania; Tanzania Industrial Research and Development Organization (TIRDO), P. O. Box 23235 Dar es salaam, Tanzania; Amref Health Africa in Tanzania, P O Box 2773 Dar es Salaam, Tanzania

**Keywords:** COVID-19, Knowledge, Attitude, Practice, Tanzania

## Abstract

**Background:** The Corona Virus Disease -19 (COVID-19) pandemic is a global health emergency that requires the adoption of unprecedented measures to control its rapid spread. Tanzanians’ adherence to control measures is affected by their knowledge, attitudes, and practices (KAP) towards the disease. This study was carried out to investigate knowledge, attitudes and practices towards COVID-19 among residents in Tanzania during the April – May 2020 period of the epidemic.

**Methods:** This cross-sectional study analyzes responses of self-selected Tanzanians who responded to an invitation to complete an online questionnaire. Survey Monkey tool was used to develop the questionnaire used for data collection. The survey assessed demographic characteristics of participants as well as their knowledge, attitudes, and practices toward COVID-19. A Chi-square analysis was used to compare proportions. Analysis of variance (ANOVA) was used to determine differences among age groups, whereas results were considered significant if the p-value was <0.05

**Results:** Four hundred residents completed the survey. The mean age of study participants was 32 years, and the majority was female (n= 216,54.0%). There were no significant differences in demographic variables). Participants with a bachelor’s degree or above (n= 241, 60.3%) had higher scores. Overall, 84.4% (n=338) of participants had good knowledge, which was significantly associated with education level (p=0.001). Nearly all participants (n=384, 96.0%) had confidence that COVID-19 will be eliminated. The majority of respondents (n=308, 77.0%) did not go to a crowded place in recent days. Multiple linear regression analysis showed that males, age-group 16-29 years, and education of secondary or lower (OR = 1.2, CI = 1.3–1.5) were significantly associated with lower knowledge score.

**Conclusions:** Our findings revealed good knowledge, optimistic attitudes, and appropriate practices towards preventing COVID-19 infection. Suggesting that community-based health education programs about COVID-19 is helpful and necessary to control the disease.

## INTRODUCTION

Coronavirus disease 2019 (COVID-19) is a disease caused by a novel coronavirus that was first detected in December 2019 in Wuhan, China. It is characterized by sudden onset, fever, fatigue dry cough, myalgia, and dyspnea. Data show that 10-20% of the patients develop severe cases, which are characterized by acute respiratory distress syndrome, septic shock, difficult-to-tackle metabolic acidosis, and bleeding and coagulation dysfunction [1,2]. World Health Organization (WHO) declared it a public health emergency of international concern and has called for collaborative efforts to prevent its rapid spread [5]. Although clinical data have shown the overall case fatality rate of COVID-19 ranges 2-5% worldwide, which is much lower than those of Severe Acute Respiratory Syndrome (SARS) (9.5%), MERS (34.4%), and H7N9 (39.0%), pathogens continue to emerge and spread to the population at risk. The threat of more contagious and virulent variants demands public health actions to move from purely treatment activities to preventive measures practiced by the public [1-3]. The ongoing COVID-19 pandemic has spread very quickly, and to date at the time of the survey, the virus had reached over 200 countries altogether, resulting in 236,368,875 laboratory-confirmed infections and 4,826,358deaths [4].

Tanzania is among African countries that have been affected by the COVID-19 pandemic [6]. Until April 24, 2021 government authorities announced only 284 cases of COVID-19, among them 256 were in stable condition, seven in special care, 37 recovery, and 10 deaths [7]. Dar-es-salaam City and Zanzibar Island have the highest number of cases. Other regions affected were Mwanza, Dodoma, Pwani, Kagera, Manyara, and Morogoro [8]. Several measures have been adopted to control COVID-19 transmission in Tanzania, including closing all schools and universities, observing physical distancing, prohibiting mass gatherings, isolating suspected cases, and caring for confirmed and suspected cases. Moreover, Tanzania residents were obliged to perform handwashing with soap and running water or alcohol-based hand sanitizer and urged to wear face masks outside their homes.

To effectively control COVID-19 in Tanzania., individuals’ adherence to these control measures is essential [9-10]. Reports from different outbreaks recommend that knowledge and attitudes towards infectious disease can sometimes result in a level of panic among the population and complicate endeavors to prevent the spread of the disease [9-12].

There is an urgent need to understand the public’s awareness of COVID-19 in Tanzania to facilitate outbreak management of COVID-19. This study was carried out to investigate knowledge, attitudes, and practices (KAP) towards COVID-19 among residents in Tanzania during the March –April 2020 period of the pandemic.

## MATERIAL AND METHODS

### Study design

This cross-sectional survey took place from 15^th^ - 28^th^ of April 2020.

### Sample size and selection

Participants were self-selected when they choose to answer the questionnaire. A total of 5,000 residents were assumed to actively use media connections. Calculation using statistical software gave a minimum sample of 384 [13]. To adjust for non-responders 400 residents conveniently receive the link with the Questionnaire [14].

### Data collection

Due to the infectious nature of disease transmission, online survey forms were used to collect the data. Survey Monkey tool was used to develop a link and KAP questionnaire for data collection.

The questionnaire consisting of 20 questions was were prepared by following WHO guide for KAP questionnaire development. This template was modified and adapted to the Swahili language. It was then validated and piloted according to guidelines for clinical and community management of COVID-19 by the Tanzanian Ministry of Health Community Development, Gender, Elderly and Children. Demographic variables included age, gender, and education Level. There were 17 KAP questions regarding clinical presentations, transmission routes, and control of COVID-19.These questions were answered either as Yes/No, true/false, and “I don’t know”. The total score ranged from 0 to 17, with a higher score indicating a better knowledge of COVID-19. Participants could provide only one response per question. The reliability of the knowledge, attitude, and practice questionnaires was checked and the values of Cronbach’s alpha were 0.81, 0.88, and 0.86 respectively, indicating acceptable internal consistency.

### Data analysis

Response data were recoded and analyzed using SPSS 17. Results were considered significant if the p-value was <0.05.

## RESULTS

### Socio-demographic characteristics of the study population

A total of 400 participants completed the survey. The range of the age of the participants was 18 to 75 years, and the mean (SD) was 32 (10.3) years. Women represented 54% of the participants. Majority of the participants (n=242, 60.3%) had a bachelor’s degree or above. Participant socio-demographic characteristics are given in Table 1.

**Table 1:** Sociodemographic characteristics of study participants (N=400)

| Variables | Frequency n (%) |
| --- | --- |
| **Sex** |  |
| Female | 216 (54.0) |
| **Age (years)** |  |
| 16-29 | 34 (08.5) |
| 30-49 | 247 (61.7) |
| 50-59 | 114 (28.5) |
| 60+ | 5 (01.3) |
| **Education Level** |  |
| Primary | 68 (17.0) |
| Secondary | 90 (22.5) |
| College/University | 242 (60.5) |

### Knowledge regarding COVID-19

Overall, 84.4% of the participants scored above the mean. The range of correct answers was two to ten the mean (SD) score was 9.3 (2.0), and the median was nine. A score above nine was considered a good knowledge level. 60.5% of the degree holder score above 9, Figure 1.

**Figure 1:**
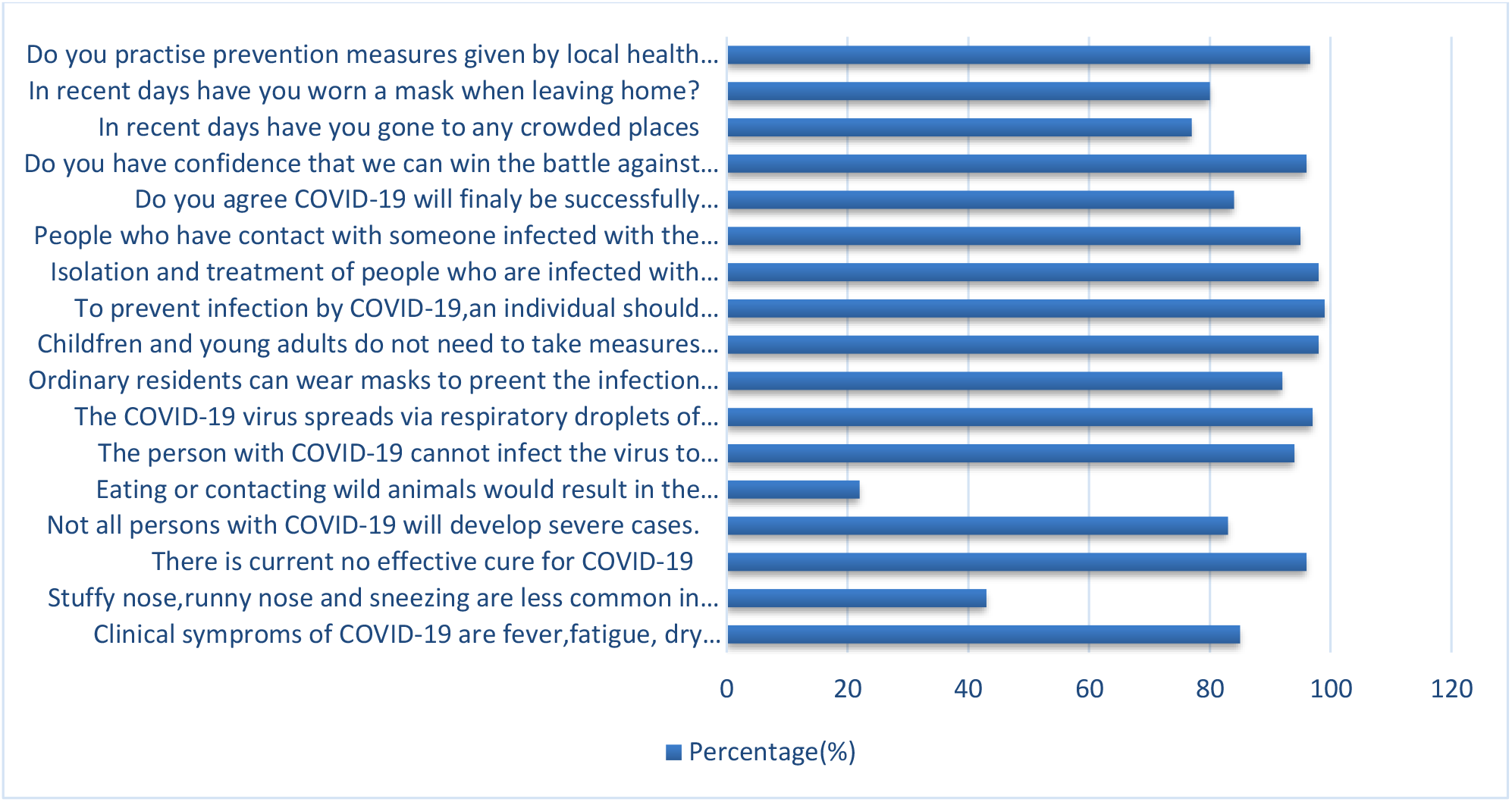
Percentage distribution of correct scores.

The range of percent of correct answers was 70.2-98.6% (Table2). The mean (SD) score was 8.7 (1.6). Results varied between genders and among age groups and education levels (P<0.001). Males, age group of 16-29 years, with a bachelor’s degree or lower were significantly associated with lower knowledge score Table 2.

**Table 2:** Distribution of mean COVID-19 knowledge by demographic variables

| Characteristics | Knowledge Level<br>n |  | Mean score<br>(SD) | P-Value |
| --- | --- | --- | --- | --- |
|  | Good | Poor |  |  |
| **Sex** |  |  |  |  |
| Male | 151 | 33 | 8.5 (2.0) | <0.001 |
| Female | 157 | 59 | 8.9 (1.3) |  |
| **Age-group (years)** |  |  |  |  |
| 16-29 | 30 | 4 | 8.(1.9) | <0.001 |
| 30-49 | 215 | 52 | 9.1(1.2) |  |
| 50+ | 90 | 36 | 8.9 (1.3) |  |
| **Education** |  |  |  |  |
| Primary | 30 | 38 | 7.7 (2.4) | <0.001 |
| Secondary | 43 | 47 | 8.8 (1.5) |  |
| College/University | 235 | 7 | 10.0 (1.2) |  |

### Attitudes and Practices Regarding COVID-19

The majority of the respondents agreed that COVID-19 would be successfully controlled (n=384, 96%). The attitude towards the final success in controlling COVID-19 had no significant differences across genders and education levels (P<0.71). The majority of the participants had not visited crowded places (n=308, 77.0%) and wore masks when going out (n=352, 80.0%) in recent days. Almost all participants (n=392, 98.0%) correctly identified that COVID-19 is transmitted through respiratory droplets and that factors such as chronic illnesses and obesity can lead to a serious morbidity (Figure 1).

## DISCUSSION

This is the first online cross-sectional study examining KAP towards COVID-19 among Tanzania residents. In this study, 84.4% of the participants had a good knowledge of COVID-19, which is comparable with a study conducted in China where more than seventy percent of study participants had good knowledge. Previous studies from different countries have identified good knowledge in infection control as a predictor of good practice [17, 18]. These studies also highlighted that major gaps in disease knowledge could result in uncertainties and non-stringent control measures [19].

Most of the participants had confidence that COVID-19 will be contained and had a certainty that we can win the fight against the disease. This attitude could have attributed to positive practice with majority reporting not visiting crowded places and wearing masks whenever they go out of their homes. Furthermore, nearly all reported adhering to preventive measures as instructed by their national health care authority. The findings are useful for policy makers to consider the need for a comprehensive specific group target health education program for COVID-19 prevention and control.

The finding of a high level of knowledge among residents is a good predictor of positive impact initiative to involve the community in a fight for COVID-19. However, the results may have been affected by the convenience sampling method used. The majority of participants (n=278, 69.5%) held a secondary degree or higher, and all participants had access to the internet. These are the privileged group who would actively seek information of this infectious disease from various channels of information, including the official website of the Ministry, and the WhatsApp account. The same study should be conducted among vulnerable populations or underprivileged communities.

The strength of this study lies in its large sample recruited during a peak of the COVID-19 outbreak. Nevertheless, compared to the population statistics, our sampling method picked middle and high economic societies, we speculate that knowledge might have been over overestimated and attitude and practice underestimated. Community based national cross-sectional study is recommended when an outbreak is over.

## CONCLUSIONS

Our findings emphasize the need to investigate the KAP towards COVID-19 among Tanzania residents of low socioeconomic status. This will encourage an optimistic attitude and maintaining safe practices. Community-based health education programs about COVID-19 are likely to be helpful and necessary approach to control the disease.

## Data Availability

Data available on request

## ACKNOWLEDGMENTS

We are grateful to all participants who took time to fill the questionnaire.

## AUTHORS DETAILS

^1^Department of Oral health, Muhimbili National Hospital-Mloganzila, Dar-Salaam, Tanzania; ^2^Tanzania Industrial Research and Development Organization (TIRDO), P. O. Box 23235 Dar es salaam, Tanzania and ^3^Amref Health Africa in Tanzania, P O Box 2773 Dar es Salaam, Tanzania

## APPENDIX 1; Questionnaire

**Knowledge, attitudes, and practices towards COVID-19 among residents during the rapid rise period of the COVID-19 outbreak**

### Demographic characteristics

1. **Age**

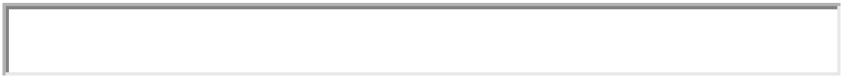
2. **Sex**

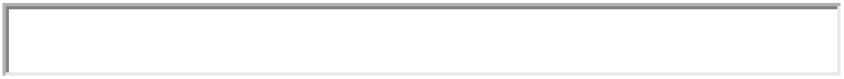
3. **Education level**

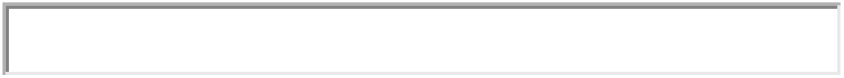

### Knowledge about Clinical characteristics of COVID-19

4. **The main clinical symptoms of COVID-19 are fever, fatigue, dry cough, and myalgia**.
  ○ True
  ○ False
  ○ I don’t know
5. **Unlike the common cold, stuffy nose, runny nose, and sneezing are less common in persons infected with the COVID-19 virus**.
  ○ True
  ○ False
  ○ I don’t know
6. **There is current and no effective cure for COVID-19 but early symptoms and supporting treatment can help most patients recover from the infections**.
  ○ True
  ○ False
  ○ I don’t know
7. **Not all persons with COVID-19 will develop severe cases. Only those who are elderly, have chronic illnesses and are obese are more likely to be severe cases**
  ○ True
  ○ False
  ○ I don’t know
8. **Eating or contacting wild animals would result in the infection by the COVID-19 virus**.
  ○ True
  ○ False
  ○ I don’t know
9. **The person with COVID-19 cannot infect the virus to others when a fever is not present**.
  ○ True
  ○ False
  ○ I don’t know
10. **The COVID-19 virus spreads via respiratory droplets of infected individuals**.
  ○ True
  ○ False
  ○ I don’t know
11. **Ordinary residents can wear masks to prevent the infection by the COVID-19 virus**.
  ○ True
  ○ False
  ○ I don’t know
12. **Children and young adults do not need to take measures to prevent the infection by the COVID-19 virus**.
  ○ True
  ○ False
  ○ I don’t know
13. **To prevent infection by COVID-19, an individual should avoid going to crowded places**.
  ○ True
  ○ False
  ○ I don’t know
14. **Isolation and treatment of people who are infected with the COVID-19 virus are an effective way to reduce the spread of the virus**.
  ○ True
  ○ False
  ○ I don’t know
15. **People who have contact with someone infected with the COVID-19 virus should be immediately isolated in a proper place**.
  ○ True
  ○ False
  ○ I don’t know
  **Attitude and Practice**
16. **Do you agree COVID-19 will finally be successfully controlled?**
  ○ Agree
  ○ Disagree
  ○ I don’t know
17. **Do you have confidence that we can win the battle against the COVID-19 virus?**
  ○ Yes
  ○ No
18. **In recent days have you gone to any crowded places?**
  ○ Yes
  ○ No
19. **In recent days have you worn a mask when leaving home?**
  ○ Yes
  ○ No
20. **Do you practice prevention measures given by local health care authorities?**
  ○ Yes
  ○ No

***Questionnaire: KISWAHILI***

**Uelewa, mtazamo, na mienendo ya wahazi wa kuhusu kirusi cha COVID-19 katika kipindi hiki cha mlipuko**

**Taarifa za mshikiri**

1. **Umri**

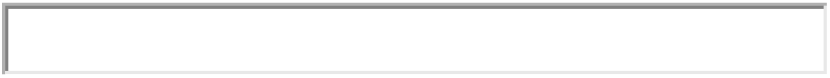
2. **Jinsia**

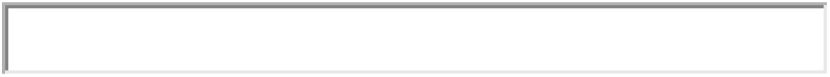
3. **Elimu**

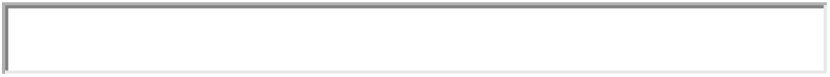 **Uelewa**
4. **Dalili kuu za mgonjwa mwenye COVID-19 ni joto kali, uchovu, kukohoa na maumivu ya misuli**.
  ○ Ndio
  ○ Hapana
  ○ Sijui
5. **Tofauti na mafua ya kawaida mgonjwa wa kirusi cha COVID-19 hapati mafua ya kutiririka, pua kuziba wala kupiga chafya**.
  ○ Ndio
  ○ Hapana
  ○ Sijui
6. **Hakuna dawa ya kutibu COVID-19 isipokua kugundua mapema hali ya maambukizi na kupata matibabu elekezi kunaweza kusaidia wagonjwa wengi kupona**.
  ○ Ndio
  ○ Hapana
  ○ Sijui
7. **Sio watu wote walio na COVID-19 watakua na kesi kali. Ila wale ambao ni wazee, wana magonjwa sugu na wenye uzito kupita kiasi wana uwezekano mkubwa wa kuwa kesi kali**
  ○ Ndio
  ○ Hapana
  ○ Sijui
8. **Kula au kugusa wanyama wa porini kunawezasababisha kuambukizwa na virusi vya COVID-19**.
  ○ Ndio
  ○ Hapana
  ○ Sijui
9. **Mtu mwenye maambukizi ya kirusi cha COVID-19 hawezi kuambukiza wengine wakati hana homa**.
  ○ Ndiyo
  ○ Hapana
  ○ Sijui
10. **Virusi vya COVID-19 huenea kupitia matone ya kupumua ya watu walioambukizwa**
  ○ Ndiyo
  ○ Hapana
  ○ Sijui
  **Mienendo**
11. **Wakazi wa kawaida wanaweza kuvaa masks kuzuia maambukizi ya kirusi cha COVID-19**
  ○ Ndiyo
  ○ Hapana
  ○ Sijui
12. **Sio lazima kwa watoto na vijana kuchukua hatua za kuzuia kuambukizwa na kirusi cha COVID-19**
  ○ Ndiyo
  ○ Hapana
  ○ Sijui
13. **Ili kuzuia kuambukizwa na COVID-19, ni vyema mtu aepuke kwenda kwenye sehemu zenye mikusanyiko**.
  ○ Ndio
  ○ Hapana
  ○ Sijui
14. **Kujitenga na kupata matibabu elekezi ya watu ambao wameambukizwa na virusi vya COVID-19 ni njia bora ya kupunguza kuenea kwa virusi**.
  ○ Ndiyo
  ○ Hapana
  ○ Sijui
15. **Watu ambao wamekutana na mtu aliyeambukizwa na virusi vya COVID-19 wanapaswa kujitenga mara moja kwa siku zisizopungua kumi na nne**.
  ○ Ndiyo
  ○ Hapana
  ○ Sijui
16. **Je! Unakubali COVID-19 hatimaye itadhibitiwa kwa mafanikio?**
  ○ Nakubali
  ○ Sikubali
  ○ Sijui
17. **Je! una imani kwamba Tunaweza kushinda vita dhidi ya virusi vya COVID-19**.
  ○ Ndiyo
  ○ Hapana
  ○ Sijui
  **Tabia**
18. **Je! Katika siku za hivi karibuni umeenda kwenye sehemu yoyote yenye mkusanyiko?**
  ○ Ndiyo
  ○ Hapana
  ○ Sijui
19. **Katika siku za hivi karibuni umevaa mask wakati wa kuondoka nyumbani?**
  ○ Ndiyo
  ○ Hapana
  ○ Sijui
20. **Je unafuata miongozo unayopewa na mamlaka za afya eneo unaloishi dhidi ya maambukizi ya COVID-19?**
  ○ Ndiyo
  ○ Hapana

